# Empirical antibacterial use, escalation, microbiological profile, and outcomes in febrile neutropenia: An ambispective observational study

**DOI:** 10.64898/2026.09.22.26363666

**Authors:** Trivedi Satish Girish, Suman Kumar, SK Singh

**Affiliations:** Department of Internal Medicine, Armed Forces Medical College, Pune - 411 040, Maharashtra, India

**Keywords:** Chemotherapy-Induced Febrile Neutropenia, Anti-Bacterial Agents, Drug Prescriptions, Drug Resistance, Bacterial, Treatment Outcome

## Abstract

**Background:** Febrile neutropenia requires rapid empirical antibiotic therapy, with regimen selection shaped by local prescribing practice and pathogen patterns. We described starting treatment, later drug changes, microbiological findings, and inpatient outcomes.

**Methods:** This ambispective cohort comprised 96 episodes from 90 patients admitted to a tertiary-level teaching hospital. Treatment and outcome analyses used each episode as the unit of observation. Recorded variables included the starting antibiotics, later regimen modification, microbiology, hospitalization length, invasive fungal infection (IFI) as captured in study records, and disposition. Comparisons based on escalation status were exploratory.

**Results:** Cefoperazone-sulbactam appeared in 62/96 initial regimens (64.6%), amikacin in 42 (43.8%), teicoplanin in 28 (29.2%), and meropenem in 14 (14.6%). Eighteen episodes (18.8%) required a change that broadened or substituted antibacterial therapy. Among 31 microbiologically documented infections, 22 (71.0%) yielded Gram-negative organisms and 9 (29.0%) were polymicrobial. Median length of admission was 18.5 days in the escalation group and 9 days in the remainder (p<0.001). Mortality was 27.8% (5/18) with escalation and 5.1% (4/78) without it (p=0.011), with nine inhospital deaths overall (9.4%).

**Conclusion:** Antibacterial management differed within this cohort, with treatment escalation required in 18 of 96 episodes. Gram-negative organisms comprised most microbiologically confirmed infections. Escalation marked a clinically complex course with prolonged hospitalization and greater observed mortality; the observational design does not establish comparative efficacy or causation.

## Introduction

Febrile neutropenia (FN) is a major complication of anticancer therapy. Prompt assessment and empirical antibacterial treatment are central to early management. Infection risk is shaped by the depth and duration of neutropenia, the underlying malignancy, treatment intensity, comorbidity, previous antimicrobial exposure, and institutional microbial ecology. Guidelines recommend broad-spectrum treatment at presentation, followed by review as bedside findings, cultures, and other microbiological data become available.1

Microbial epidemiology differs between institutions and evolves over time. Indian tertiary-center studies document a substantial burden of Gram-negative infection together with antimicrobial resistance, supporting locally generated surveillance when empirical policies are developed.2-5

Cefoperazone-sulbactam is one empirical option used in several Asian settings. A systematic review of comparative studies found comparable treatment success and mortality between cefoperazone-sulbactam and alternative regimens, with selection still guided by local susceptibility patterns and individual clinical risk.6 Evidence is less complete for the course after first-line treatment, particularly why antibiotics are broadened or changed and how those decisions relate to microbiology and clinical outcomes.

We examined how empirical antibacterial prescribing evolved during hospitalization, from the first regimen to subsequent changes. We also described the organism profile, later drug exposure, duration of admission, and in-hospital mortality.

## Methods

### Study design and participants

This ambispective project combined review of existing charts with prospective enrollment at a tertiary-care hospital in western India from 23 September 2024 to 23 March 2026. Historical records supplied one component, whereas eligible participants entering the prospective arm were enrolled consecutively. Treating teams made all clinical decisions independent of the study. A qualifying FN event was the principal analytical unit.

Eligibility required age ≥12 years and hospital admission for FN associated with a hematologic or solid-organ malignancy. Outpatient presentations and fever attributed to noninfectious causes were excluded. A patient could contribute another qualifying event during a separate treatment cycle.

Sample size was derived from n = Z^2^p(1-p)/d^2^ using 95% confidence, p=0.50, and absolute precision 0.10; the minimum calculated requirement was 96 observations.

### Definitions

FN was defined as a single oral temperature >38.3°C or a temperature ≥38.0°C sustained for more than 1 hour, together with an absolute neutrophil count <500 cells/µL or an anticipated fall below that level within 48 hours.1

Microbiologically documented infection (MDI) required recovery of a clinically relevant pathogen from an appropriate specimen in a compatible clinical setting. Clinically documented infection (CDI) denoted an identifiable clinical or radiological focus without microbiological confirmation. Observations outside both definitions formed a third category.

First empirical therapy referred to antibiotics started before definitive microbiological information became available. Escalation meant adding or replacing an antibacterial agent in response to persistent fever or inadequate response, an emerging infective focus, microbiological results, or clinical deterioration. Because 72-hour response was captured only inconsistently as a separate variable, non-escalation was treated strictly as the absence of a regimen change rather than as a surrogate for documented response.

### Data collection and outcomes

Structured study records plus the anonymized master file supplied demographics, malignancy type, chemotherapy, neutropenia duration, antibiotic exposure, regimen changes, reasons for escalation, microbiology, clinical focus, recorded IFI, antifungal therapy, length of admission, and final disposition.

Primary endpoints were the initial antibiotic pattern and the need for later escalation. Secondary measures covered the trigger for modification, drugs given afterward, infection classification and organism spectrum, IFI, duration of hospitalization, and final outcome.

Microbiology was assessed per FN event. Recovery of multiple clinically relevant organisms retained a polymicrobial classification, allowing organism-specific percentages to overlap. Given incomplete susceptibility testing and phenotype documentation, resistance prevalence was left for future analysis with more complete data.

IFI status was taken directly from study records.

### Statistical analysis

Demographic characteristics were summarized once per patient. Treatment, infection, and outcome variables used individual FN events as observations. Continuous data are presented as mean±standard deviation or median (interquartile range), and categorical data as n (%); missing values were not imputed.

Exploratory comparisons used the Mann-Whitney U test for continuous variables and Fisher exact test for categorical variables when appropriate. Selected binary outcomes were summarized with crude risk ratios and 95% confidence intervals. A sensitivity analysis limited the dataset to each patient’s first qualifying event to assess the influence of repeated observations. Tests were two-sided with p<0.05 as the significance threshold. Inferential results were treated as exploratory because sample-size planning targeted precision around a single proportion instead of formal hypothesis testing between groups.

### Ethical considerations

The Institutional Ethics Committee approved the study (IEC/INT MED/136/2024; 18 April 2024). Prospective participants provided written informed consent; archived records were reviewed under a consent waiver. Personal identifiers were removed before analysis.

## Results

### Study population

Ninety patients contributed 96 qualifying FN events. Age was available for 87 individuals: mean 49.3±17.0 years, median 53 years (IQR 36-61.5). There were 58 men (64.4%). Hematologic malignancy affected 67/90 patients (74.4%); the remaining 23 (25.6%) had a solid malignancy.

Thirty-one of 96 observations (32.3%) met MDI criteria, 21 (21.9%) met CDI criteria, and 44 (45.8%) belonged to neither category. Neutropenia duration was recorded in 95 observations, with a median of 5 days (IQR 3-7); 17/95 (17.9%) exceeded 7 days. Hospitalization lasted a median 9 days (IQR 6-15.25; range 2-63).

### Initial empirical antibacterial therapy

The most common initial exposure was cefoperazone-sulbactam (62/96; 64.6%). Amikacin appeared in 42 events (43.8%), teicoplanin in 28 (29.2%), meropenem in 14 (14.6%), piperacillin-tazobactam in 12 (12.5%), levofloxacin in 3 (3.1%), and vancomycin in 1 (1.0%). Combination regimens were common. Ceftriaxone, amoxicillin-clavulanate, and clindamycin each occurred once among other starting regimens.

### Antibacterial escalation

Antibacterial escalation occurred in 18/96 events (18.8%). Persistent or non-resolving fever alone explained 7/18 (38.9%). Eight modifications (44.4%) followed a newly identified clinical focus, microbiological information, or both; one event (5.6%) combined persistent fever with a new focus. Two cases (11.1%) lacked a clearly documented reason.

Teicoplanin appeared in 13/18 escalated courses (72.2%) and meropenem in 11/18 (61.1%). Additional agents were tigecycline, colistin, piperacillin-tazobactam, aztreonam, linezolid, ceftazidime-avibactam, amikacin, levofloxacin, and vancomycin. The master file contained one field for both second- and third-line therapy rather than separate entries. Accordingly, these agents are reported collectively as post-escalation exposure.

### Microbiology with clinical outcomes

Pathogens were documented in 31 FN events. Gram-negative organisms occurred in 22/31 (71.0%), Grampositive organisms in 10/31 (32.3%), and 9/31 (29.0%) were polymicrobial. Klebsiella species were identified in 11 MDI events (35.5%), Escherichia coli in 7 (22.6%), Staphylococcus aureus in 6 (19.4%), Enterococcus species in 4 (12.9%), and Acinetobacter and Pseudomonas species in 3 each (9.7%). IFI appeared in 12/96 observations (12.5%).

Disposition was discharge in 82/96 events (85.4%), death during admission in 9 (9.4%), and discharge against medical advice in 5 (5.2%).

Median stay was 18.5 days in escalated events and 9 days otherwise (p<0.001). Median neutropenia duration was 7 versus 5 days (p=0.008), and MDI was present in 55.6% versus 26.9% (p=0.026).

Mortality was 27.8% (5/18) after escalation and 5.1% (4/78) without it (Fisher exact p=0.011), corresponding to a crude risk ratio of 5.42 (95% CI 1.61-18.18). IFI was recorded in 3/18 and 9/78, respectively (p=0.693). Restricting the analysis to each patient’s first event yielded mortality of 26.7% (4/15) versus 5.3% (4/75); median admission duration remained approximately 18 versus 9 days.

## Discussion

Three findings stand out. Prescribing at presentation varied, although cefoperazone-sulbactam was the most frequent component. Fewer than one in five FN events required a later regimen change. Gram-negative organisms accounted for most microbiologically confirmed infections, and patients whose therapy was escalated tended to have longer admissions and more deaths.

Combination therapy was frequent at presentation. Indian reports likewise describe wide variation in empirical regimens and emphasize institution-level microbiological surveillance.2-5 Babu et al. found a sizeable Gramnegative bloodstream-infection burden alongside temporal susceptibility shifts that influenced local empirical policy.2 Ghosh et al. reported a similar Gram-negative predominance in high-risk FN and used cefoperazonesulbactam plus amikacin as first-line treatment.3

Because clinicians chose regimens rather than allocating them for comparison, relative effectiveness cannot be inferred. The 81.3% non-escalation rate also cannot be attributed to cefoperazone-sulbactam, which appeared in 64.6% of initial regimens. Published comparisons suggest similar clinical success and mortality across cefoperazone-sulbactam and comparator regimens, with antibiotic choice still guided by patient risk and local susceptibility.6

Teicoplanin was included at presentation in 29.2% of observations. The dataset lacked consistent indicationlevel detail, preventing classification of individual glycopeptide prescriptions as appropriate or inappropriate. This frequency is better treated as a prompt for a focused prescribing audit.

Persistent fever and newly emerging clinical or microbiological information were the main reasons for modifying therapy; meropenem and teicoplanin were often introduced at that point. Such changes usually occur in patients whose clinical course is already difficult. We interpret this association with death and prolonged admission as reflecting illness severity and treatment complexity, not an effect of the regimen change itself.

Microbiological confirmation occurred in roughly one-third of FN observations, and Gram-negative organisms formed the largest group, in keeping with Indian cohorts.2-5 Nine MDI events were polymicrobial. An eventbased summary retains more information from these records than a forced single-isolate assignment. Incomplete susceptibility information prevented reliable prevalence estimates for extended-spectrum betalactamase production, carbapenem resistance, methicillin-resistant S. aureus, or vancomycin-resistant enterococci. Future surveillance would be stronger with prospective capture of isolate-level susceptibility results.

Recorded IFI affected 12.5% of observations. EORTC/MSGERC classification depends on specified host, clinical, and mycological evidence, with more stringent requirements for proven disease.7 Those criterionlevel details were not uniformly recoverable, so the original possible/probable/proven labels were omitted from inferential analysis.

Nine patients died in hospital (9.4%). Mortality and admission duration were both higher in the escalation group. This pattern is more consistent with escalation occurring in patients who already have a difficult clinical course than with escalation causing the adverse outcome. Results were similar in the first-event analysis, making repeated observations an unlikely sole explanation.

Interpretation is constrained by the single-center setting, uneven completeness of archived documentation, and nonrandom treatment selection. Formal 72-hour response data and complete susceptibility results were missing for some observations, and several patients contributed more than one event. Sample-size planning was intended for estimation rather than outcome modeling; only nine deaths occurred, which precluded a reliable multivariable mortality model. The records did not permit uniform contemporary re-adjudication of IFI.

## Conclusion

Hospital prescribing was heterogeneous. Cefoperazone-sulbactam was the most common starting agent, while escalation occurred in just under one-fifth of FN events. Gram-negative pathogens accounted for most MDI, and some infections were polymicrobial. Patients who required broader or substitute therapy had longer admissions and higher observed mortality, a pattern we attribute to clinical complexity rather than to a demonstrated treatment effect. More consistent prospective recording of prescribing indications and susceptibility results would strengthen stewardship review and future protocol assessment.

**Table 1.** Baseline demographic and clinical characteristics.

| Characteristic | Value |
| --- | --- |
| Unique patients, n | 90 |
| Febrile-neutropenia episodes, n | 96 |
| Age available, n | 87 |
| Age, years - mean ± SD | 49.3 ± 17.0 |
| Age, years - median (IQR) | 53 (36-61.5) |
| Male sex, n (%) | 58/90 (64.4) |
| Hematologic malignancy, n (%) | 67/90 (74.4) |
| Solid malignancy, n (%) | 23/90 (25.6) |
| Microbiologically documented infection, n (%) | 31/96 (32.3) |
| Clinically documented infection, n (%) | 21/96 (21.9) |
| Neither MDI nor CDI, n (%) | 44/96 (45.8) |
| Neutropenia duration available, n | 95 |
| Neutropenia duration, days - median (IQR) | 5 (3-7) |
| Neutropenia >7 days, n (%) | 17/95 (17.9) |
| Hospital stay, days - median (IQR) | 9 (6-15.25) |
| Hospital stay, days - range | 2-63 |

*SD: standard deviation. IQR: interquartile range. MDI: microbiologically documented infection. CDI: clinically documented infection. Demographic variables are summarized per unique patient; episode characteristics are summarized per FN episode.*

**Table 2.** Initial empirical antibacterial exposure and treatment escalation (n = 96 episodes).

| Measure | n/N (%) |
| --- | --- |
| **Initial empirical antibacterial exposure** |  |
| Cefoperazone-sulbactam | 62/96 (64.6) |
| Amikacin | 42/96 (43.8) |
| Teicoplanin | 28/96 (29.2) |
| Meropenem | 14/96 (14.6) |
| Piperacillin-tazobactam | 12/96 (12.5) |
| Levofloxacin | 3/96 (3.1) |
| Vancomycin | 1/96 (1.0) |
| Ceftriaxone | 1/96 (1.0) |
| Amoxicillin-clavulanate | 1/96 (1.0) |
| Clindamycin | 1/96 (1.0) |
| Antibacterial escalation | 18/96 (18.8) |
| No antibacterial escalation | 78/96 (81.3) |
| **Reason for escalation (n = 18)** |  |
| Persistent/non-resolving fever alone | 7/18 (38.9) |
| New clinical focus, microbiological information, or both | 8/18 (44.4) |
| Persistent fever plus new focus | 1/18 (5.6) |
| Reason unclear/not documented | 2/18 (11.1) |
| Post-escalation teicoplanin exposure | 13/18 (72.2) |
| Post-escalation meropenem exposure | 11/18 (61.1) |

*Initial antibiotic counts are not mutually exclusive because combination regimens were common. Other post-escalation agents recorded were tigecycline, colistin, piperacillin-tazobactam, aztreonam, linezolid, ceftazidime-avibactam, amikacin, levofloxacin, and vancomycin; source records did not separate second- and third-line exposure into distinct fields.*

**Table 3.** Microbiological profile and source-recorded invasive fungal infection.

| Measure | n/N (%) |
| --- | --- |
| Microbiologically documented infection (MDI) | 31/96 (32.3) |

| Among MDI episodes (n = 31) |  |
| --- | --- |
| Gram-negative organisms | 22/31 (71.0) |
| Gram-positive organisms | 10/31 (32.3) |
| Polymicrobial infection | 9/31 (29.0) |
| Klebsiella species | 11/31 (35.5) |
| Escherichia coli | 7/31 (22.6) |
| Staphylococcus aureus | 6/31 (19.4) |
| Enterococcus species | 4/31 (12.9) |
| Acinetobacter species | 3/31 (9.7) |
| Pseudomonas species | 3/31 (9.7) |
| Source-recorded invasive fungal infection (IFI) | 12/96 (12.5) |

*Organism-specific percentages may overlap because 9 of 31 MDI episodes were polymicrobial. Resistance prevalence was not estimated because susceptibility and phenotype documentation were incomplete. IFI status is reported as captured in the study records.*

**Table 4.** Clinical outcomes and exploratory comparison according to antibacterial escalation.

| Outcome | Escalation (n = 18) | No escalation (n = 78) | p value | Effect estimate |
| --- | --- | --- | --- | --- |
| Hospital stay, median days | 18.5 | 9 | <0.001 | - |
| Neutropenia duration, median days | 7 | 5 | 0.008 | - |
| Microbiologically documented infection | 10/18 (55.6%) | 21/78 (26.9%) | 0.026 | - |
| In-hospital mortality | 5/18 (27.8%) | 4/78 (5.1%) | 0.011 | RR 5.42 (95% CI 1.61-18.18) |
| Source-recorded IFI | 3/18 (16.7%) | 9/78 (11.5%) | 0.693 | - |

**Figure 1.**
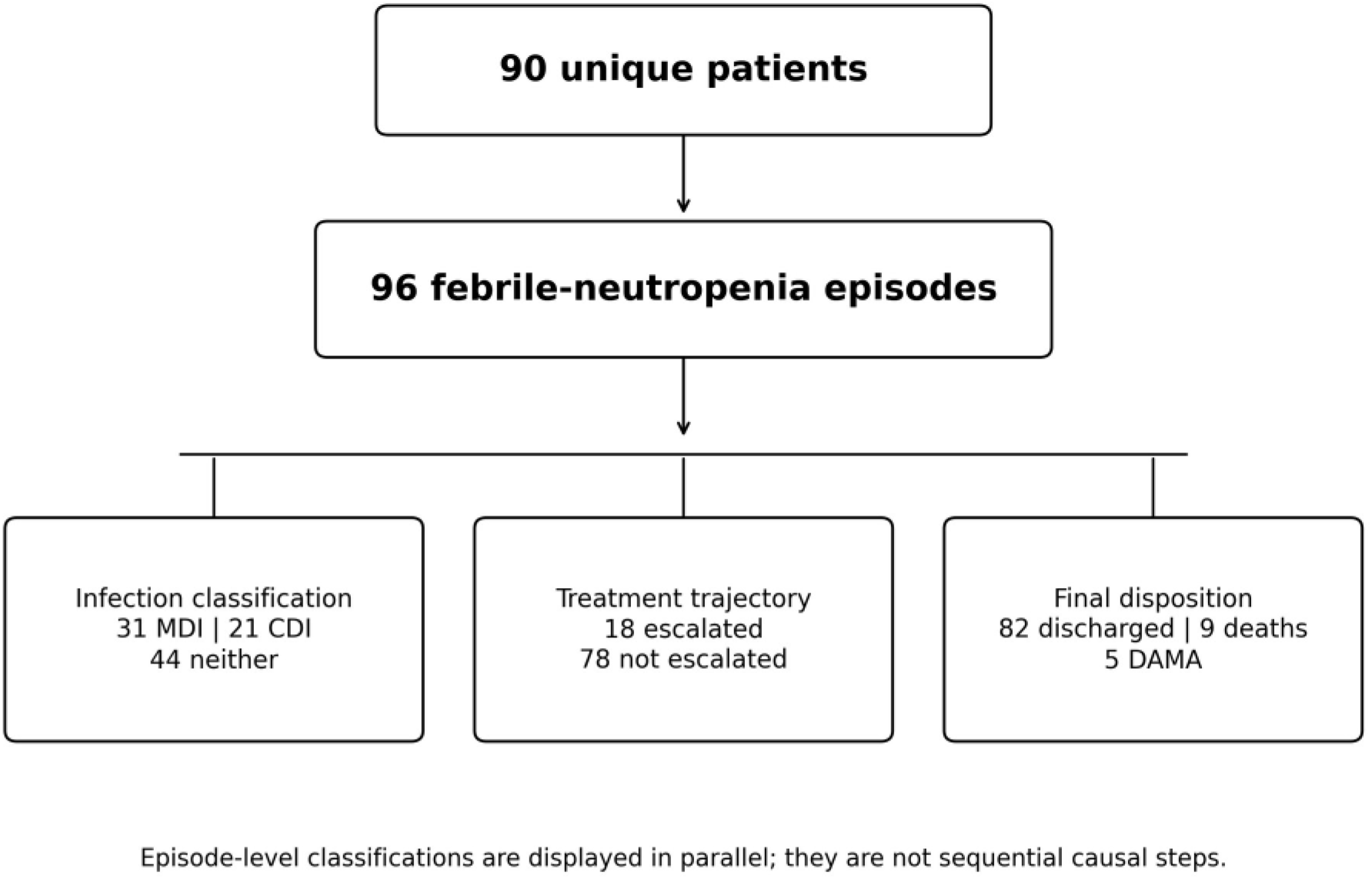
Participant and episode flow. Ninety unique patients contributed 96 febrile-neutropenia episodes. Episode-level classifications are displayed in parallel: 31 episodes were microbiologically documented infections, 21 were clinically documented infections, and 44 met neither category; 18 episodes underwent antibacterial escalation and 78 did not; final disposition was discharge in 82 episodes, in-hospital death in 9, and discharge against medical advice in 5. The parallel branches are descriptive classifications and do not represent sequential causal steps.

**Figure 2.**
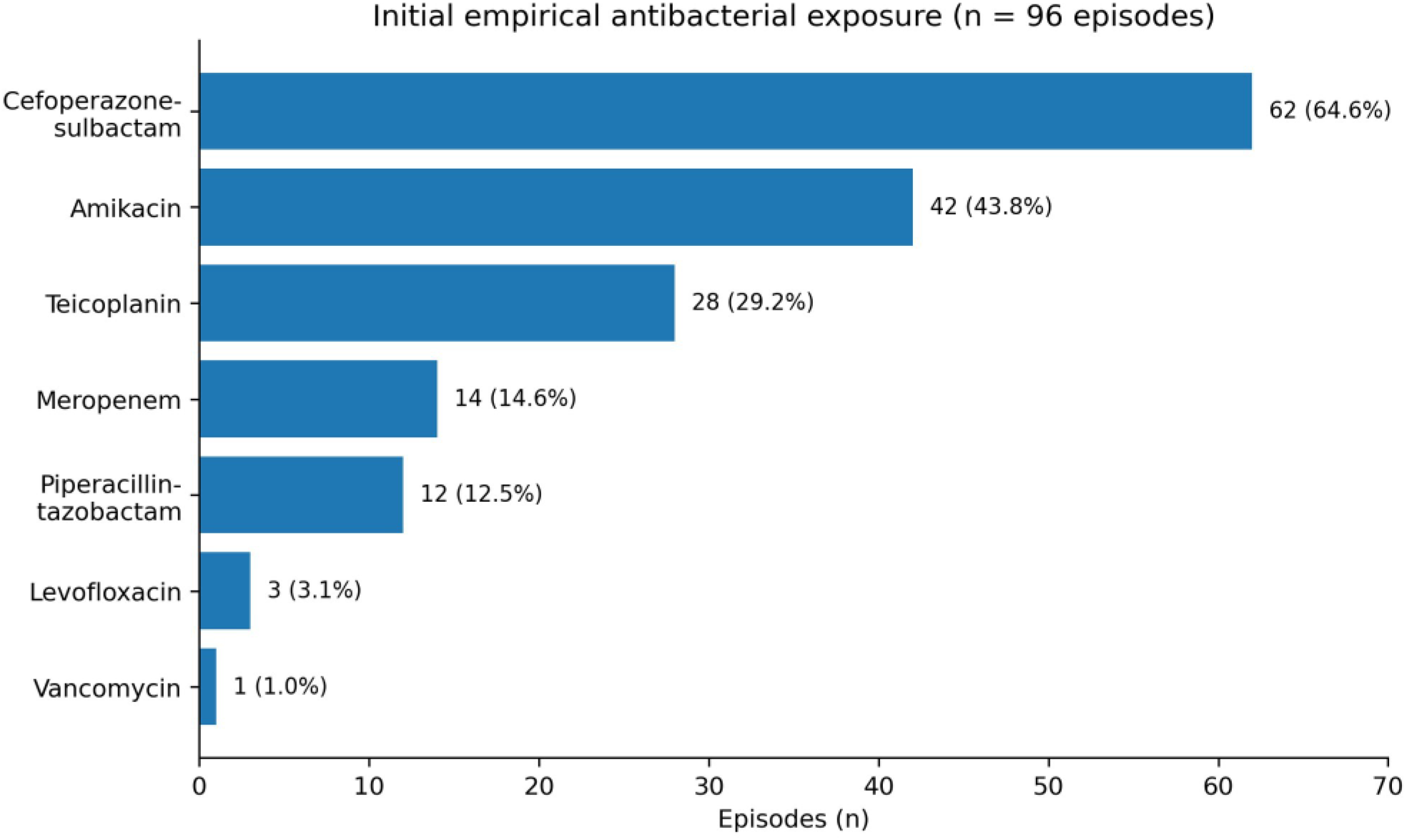
Initial empirical antibacterial exposure across 96 febrile-neutropenia episodes. Cefoperazone-sulbactam was recorded in 62 episodes (64.6%), amikacin in 42 (43.8%), teicoplanin in 28 (29.2%), meropenem in 14 (14.6%), piperacillin-tazobactam in 12 (12.5%), levofloxacin in 3 (3.1%), and vancomycin in 1 (1.0%). Because combination regimens were common, counts are not mutually exclusive.

**Figure 3.**
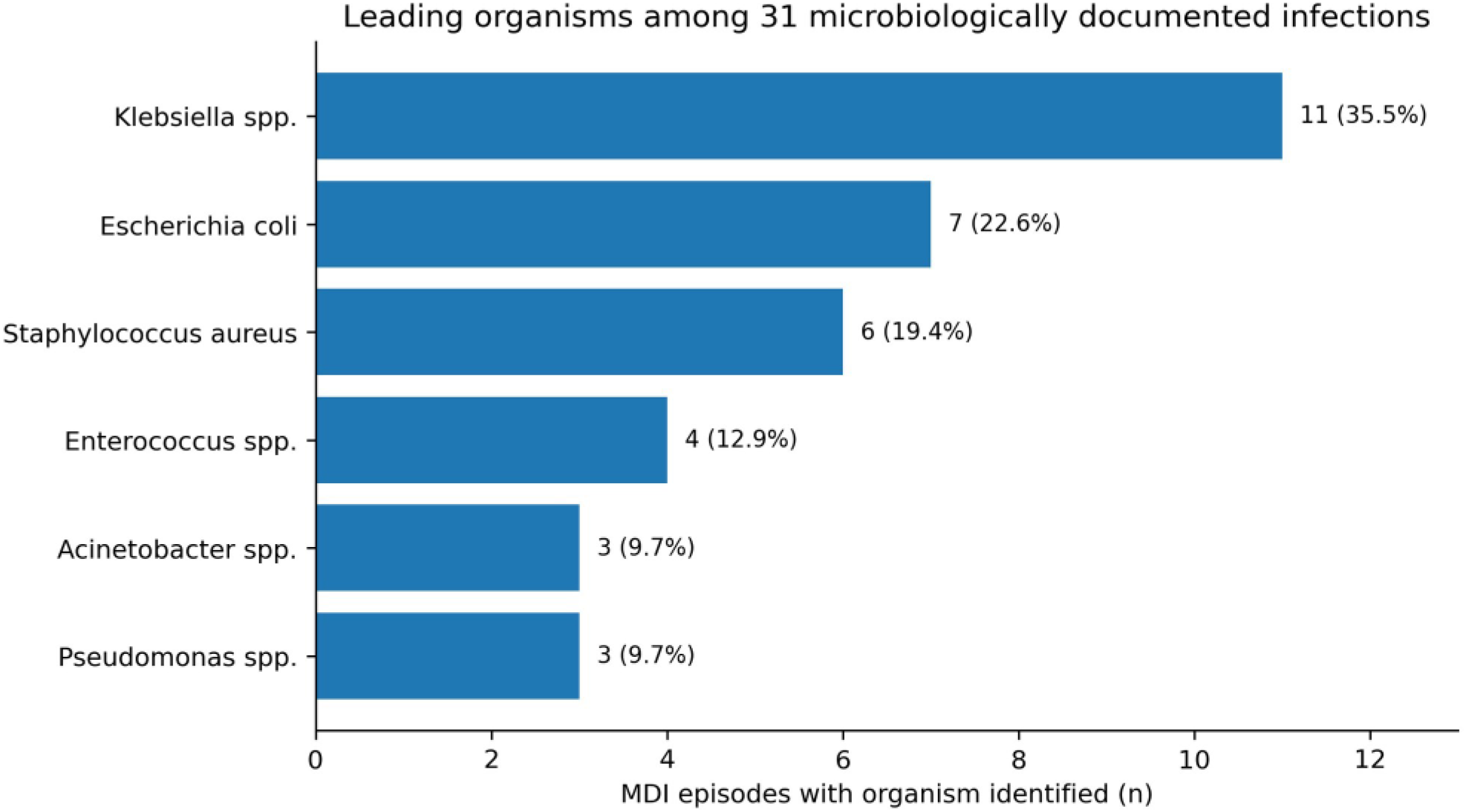
Leading organisms among 31 microbiologically documented infection episodes. Klebsiella species were identified in 11 episodes (35.5%), Escherichia coli in 7 (22.6%), Staphylococcus aureus in 6 (19.4%), Enterococcus species in 4 (12.9%), and Acinetobacter and Pseudomonas species in 3 each (9.7%). Organism counts are not mutually exclusive because 9 of 31 MDI episodes were polymicrobial.

## Data Availability

All data produced in the present study are available upon reasonable request to the authors.

## Declarations

### Ethics approval and informed consent

The study was approved by the Institutional Ethics Committee (IEC/INT MED/136/2024; 18 April 2024). Written informed consent was obtained for the prospective component; waiver of consent was granted for retrospective record review. Personal identifiers were removed before analysis.

### Data availability

The de-identified data supporting the findings of this study are available from the corresponding author on reasonable request, subject to institutional and ethical approval.

### Reporting guideline

This observational study is reported in accordance with the STROBE statement.

## Acknowledgements

The authors thank the Department of Internal Medicine, Armed Forces Medical College, Pune, and all patients who participated in this study.

